# Vaginal Antisepsis for Major Gynecologic Surgeries Using Chlorhexidine Gluconate versus Povidone Iodine: A Systematic Review and Meta-Analysis

**DOI:** 10.64898/2026.05.26.26353429

**Authors:** Yasmin Dias, Sabrina Lara, Amanda Godoi, Cintia Gomes, Lauren Yeager, Siobhan Sutcliffe, Jerry L. Lowder

## Abstract

**OBJECTIVE:** We performed a systematic review and meta-analysis (SRMA) of post-surgical outcomes, comparing chlorhexidine gluconate (CHG) versus povidone iodine (PI) for vaginal antisepsis of major gynecologic procedures.

**DATA SOURCES:** Ovid Medline, Embase, Scopus, Embase, Cochrane, and Clinicaltrials.gov were searched between 1986 and December 2023, for studies comparing CHG with PI for vaginal antisepsis of major gynecologic operations.

**STUDY ELIGIBILITY CRITERIA:** We included Randomized Controlled Trials (RCTs) and non-RCTs comparing CHG to PI for vaginal antisepsis of major gynecologic operations. The primary outcome was surgical site infections (SSIs) and the secondary outcome was urinary tract infections (UTIs) and vaginal irritation.

**METHODS:** Summary estimates were calculated by fixed effects models when I^2^ ≤ 25% and by random effects models when I^2^ > 25%. Statistical analysis was performed using RevMan 5.4.1. The protocol for this systematic review was registered on PROSPERO (ID CRD42022378101).

**RESULTS:** Nine studies met the inclusion criteria, four of which were randomized controlled trials (RCTs). 9538 patients were included, 4300 (45%) of whom were allocated to CHG and 5238 (55%) to PI. No statistically significant difference in SSI incidence was found for vaginal antisepsis with CHG versus PI in pooled analyses (n= 9538 patients; RR 1.20; 95% CI 0.92-1.57; I^2^ =0%). In contrast, a significantly higher risk of UTIs was observed for vaginal antisepsis with CHG than with PI (n=6061 patients; RR 1.48 95% CI 1.03-2.14; I^2^ = 0%).

**CONCLUSION:** In our SRMA, there were no significant differences in SSI risk when either CHG or PI was utilized for antiseptic vaginal preparation. Interestingly, vaginal antisepsis with PI was associated with a lower incidence of post-operative UTIs following major gynecologic surgery. Our findings support current guidelines that form of vaginal antisepsis can be used for SSI prevention. They also suggest that PI may result in fewer postoperative UTIs but further randomized studies are needed to support these findings.

**CONDENSATION PAGE:** *Tweetable statement:* Our meta-analysis found that chlorhexidine gluconate and povidone iodine are both acceptable for vaginal preparation in major GYN surgery.

*AJOG at a Glance:* a) Why was this study conducted? Although vaginal antisepsis is recommended before major gynecologic surgeries, there is no consensus over the preferred vaginal preparation solution. b) What are the key findings? Our systematic review and meta-analysis showed that the risk of surgical site infections (SSIs) after major gynecologic surgery was similar following vaginal antisepsis with chlorhexidine gluconate (CHG) and povidone iodine (PI). However, the risk of urinary tract infections (UTIs) was significantly lower following antisepsis with PI. c) What does this study add to what is already known? Our findings support current guidelines recommending use of either form of vaginal antisepsis for major gynecologic procedures that require vaginal preparation.

## Introduction

Postoperative surgical site infections (SSIs) are common complications of gynecologic surgical procedures and significant causes of surgical morbidity and mortality.^1,2^ The risk of SSI is estimated to be approximately 2% for hysterectomies^3^ and 1% for all gynecologic surgeries combined.^4,5^ These estimates are likely low, as many infections occur after hospital discharge and thus may be treated at outside healthcare facilities.^3^ SSIs are costly, contributing to approximately $3.5–10 billion in US healthcare expenditures annually.^4^

In addition to SSIs, urinary tract infections (UTIs) are common causes of morbidity after major gynecologic surgeries.^1^ Recent literature suggests that 7% to 33% of women develop a UTI after undergoing urogynecological surgery.^6,7^ These postoperative infections increase healthcare costs, antibiotic resistance, and morbidity and mortality for patients. They also remain an area of quality improvement for many hospitals and serve as a quality metric for regulatory organizations such as the Joint Commission and the Centers for Medicare and Medicaid Services.^8^

To reduce the risk of both SSIs and UTIs, the American College of Obstetricians and Gynecology (ACOG) recommends preoperative antiseptic preparation of the vaginal surgical field before vaginal surgical procedures.^9^ Currently, povidone iodine (PI) is the only approved vaginal antisepsis by the U.S Food and Drug Administration.^10^ However, ACOG has also proposed that 4% chlorhexidine gluconate (CHG) may be a safe and effective alternative, except in cases of allergy.^11^ For abdominal skin preparation at the time of abdominal hysterectomy, CHG has been shown to be more effective at reducing SSIs than PI in several studies.^11^ However, the data are less clear for vaginal surgery.

### Objective

The purpose of this meta-analysis is to systematically review and synthesize the published literature on vaginal antisepsis with CHG and PI to inform the most effective method of SSI and UTI prevention.

## Methods

This systematic review and meta-analysis (SRMA) was preceded by a written protocol registered in November in 2022 the International Prospective Register of Systematic Reviews (PROSPERO registration number CRD42022378101) and performed following the Cochrane Collaboration and the Preferred Reporting Items for Systematic Reviews, Meta-Analysis (PRISMA) statement guidelines and Observational Studies in Epidemiology (MOOSE) guidelines.^12–14^

### Search strategy

A medical librarian (LHY) searched the literature for records including the concepts of gynecologic surgeries, chlorhexidine, povidone Iodine, surgical site infections, UTIs, and vaginal irritation. The librarian created search strategies using a combination of keywords and controlled vocabulary in Embase.com 1947-, Ovid Medline 1946-, Scopus 1823-, Cochrane Central Register of Controlled Trials (CENTRAL), The Cochrane Database of Systematic Reviews (CDSR), and Clinicaltrials.gov 1997-. All search strategies were completed 12/15/2023 with no added limits and a total of 891 results were found. 347 duplicate records were deleted using Covidence.org and 2 duplicate records were detected manually resulting in a total of 541 unique citations included in the project library. Fully reproducible search strategies for each database can be found in the Appendix 1.

### Eligibility criteria

Included studies were required to meet the following eligibility criteria: 1) were randomized controlled trial (RCT) or observational in design; 2) compared CHG to PI; 3) included a major gynecologic procedure as the primary surgery; 4) utilized vaginal antisepsis; 5) were published in the English language; and 6) reported at least one clinical outcome of interest. We excluded studies that: 1) had overlapping patient populations; 2) were case report or case series in design; 3) did not present original research (i.e., commentaries, notes, or editorials); or 4) included pregnant individuals.

### Study selection and data extraction

Titles and abstracts were screened independently by two investigators (Y.D. and S.L.). After removing duplicate studies and abstracts that did not meet eligibility criteria, we examined the full texts of the remaining studies for eligibility. Disagreements were resolved by consensus. If multiple publications were based on the same study population, we included the study with the most comprehensive data and longest follow-up. The study selection process is shown in Figure 1. Two investigators (Y.D. and S.L.) extracted data independently from selected studies, with disagreements resolved by consensus. Extracted data included baseline characteristics reported in Table 1 and outcomes. Authors of studies were directly contacted in the event of missing information.

### Assessment of risk of bias

An objective assessment of the methodologic quality of each study was performed using the Cochrane Handbook of Systematic Reviews for evaluating randomization, concealment, blinding, intention-to-treat, baseline comparisons, concomitant interventions, and completeness of follow-up. Each study received a score of high, low, or unclear risk of bias in each domain. Risk of bias assessment was performed independently by two authors (A.G. and C.G.). Disagreements were resolved by consensus with a third reviewer (Y.D.).

RCTs were assessed using the revised tool to assess risk of bias in randomized trials (RoB 2).^15^ For non-randomized studies, we used the tool for assessing risk of bias in non-randomized studies of interventions (ROBINS-I).^16^ All analyses were conducted with an intention-to-treat approach.

### Data synthesis

Data analysis was performed using Review Manager version 5.4.1 (Nordic Cochrane Centre, The Cochrane Collaboration, Copenhagen, Denmark). Heterogeneity was measured using the Cochrane Q test and I-squared (Higgins I^2^) test. Values considered significant for heterogeneity included a p-value inferior to 0.10 and I^2^ > 25%. A fixed-effects model was used to calculate pooled relative risks (RRs) and 95% confidence intervals (CIs) for endpoints with *I*^2^ ≤ 25% (low heterogeneity). A DerSimonian and Laird random-effects model was used for pooled outcomes with high heterogeneity. We also performed funnel plot analysis for appraisal of publication bias.

### Sensitivity and subgroup analyses

A priori, we planned subgroup analyses restricted to: 1) RCTs to produce pooled estimates less susceptible to confounding; and 2) published articles to produce pooled estimates less susceptible to errors that might be detected by peer review. In addition, we performed funnel plot analysis for appraisal of potential small study effects (publication bias).

## Results

### Study Selection

As shown in Figure 1, 890 studies were identified overall. Duplicate studies and those ineligible by title and abstract review were excluded, leaving 27 articles. These were fully reviewed according to eligibility criteria; the main reasons for exclusion were lack of a control group or intervention of interest, pregnant population, and lack of outcomes of interest.

### Study Characteristics

Nine studies met all criteria and were included, comprising 9538 patients from four RCTs ^17–20^ and five observational studies of which three were pre-post cross-sectional studies and two were retrospective cohorts.^21–25^ All studies were published in the United States of America. Primary and secondary outcomes were pre-specified, where the primary outcome was SSIs for all the studies. Secondary outcomes were comprised of UTIs and vaginal irritation (symptoms such as dryness and itching). Al-Niaimi et al. monitored postoperative vaginal irritation by applying a numeric visual analog scale of vaginal irritation reported by the patients on the first day after surgery.^21^ Rastogi et al,^18^ also reported their results based on a standard instrument PRO-CTCAE (Patient-Reported Outcomes version of the Common Terminology Criteria for Adverse Events) created by the National Institute of Health and National Cancer Institute.^26^ Rockefeller reported their results based on a 5-point Likert scare on postoperative day 1.^20^ Lastly, UTI was reported in three studies and considered with positive cultures.^19,20,22^ Positive cultures if the bacterial count was ≥5,000 colony-forming units (CFUs) based on previous studies.^17,27^ Most studies included patients undergoing hysterectomies, whereas one study included patients undergoing urogynecology surgeries, such as ligament suspension, colporrhaphy, cystoscopy, colporrhaphy.^20^ Follow up periods ranged from 24 hours to 6 weeks postoperative.

### Risk of bias

Figure 2 present the individual quality assessment of RCTs and observational studies included in the meta-analysis.^12,15,28^ All randomized studies had an appropriate randomization process and were classified as “low” risk of bias. The observational studies were classified as serious or moderate risk of bias due to lack of control for important potential confounders such as patient clinical and perioperative variables.^21,22,29^ As shown in Supplementary Figure 1, the funnel plot showed a symmetrical distribution of similar-weight studies with convergence towards the pooled treatment effect size as weights increased. There was no evidence of publication bias.

### Synthesis of Results

Figures 3 and 4 present the pooled analysis of all studies. No significant differences were found between groups that received CHG versus PI antisepsis for the outcomes of SSI (RR 1.20; 95% CI 0.92 - 1.57; *I*^2^ = 0%) or vaginal irritation (RR 1.23; 95% CI 0.28 - 5.40; *I*^2^ = 69%). In contrast, patients receiving vaginal antisepsis with CHG had significantly increased risk of UTIs compared to those that received antisepsis with PI (RR 1.48; 95% CI 1.03 - 2.14; p = 0.03; *I*^2^ = 0%). (Figure 5)

In sub-analyses limited to RCTs, too few participants were included and too few infections were observed to provide stable pooled estimates (n=119 patients from the 1 RCT that observed a total of 3 infections, RR 0.48, 95% CI 0.04-5.10 for SSIs; and n=191 patients from 2 RCTs; RR 1.39, 95% CI 0.64-3.05; *I*^2^ =0% for UTIs). (Figures 6 and 7) Analyses limited to published studies yielded similar estimates as those including all studies for SSIs (n=8305 patients; RR 1.27; 95% CI 0.94-1.72; *I*^2^ =0% for SSIs). (Figure 8) All studies that evaluated UTIs were published as peer-reviewed manuscripts.

## Comment

### Principal Findings

This SRMA observed no significant differences in the primary outcome of SSI risk between patients who received either CHG or PI as a vaginal preparation before major gynecologic procedures. However, a higher risk of UTIs was observed in patients who underwent vaginal preparation with CHG than with PI. Additionally, no significant differences were observed in vaginal irritation between groups. Based on our findings, CHG and PI appear to be equally effective for vaginal antisepsis prior to major gynecologic surgery; however, further investigation into PI may be warranted for its potential to lower risk of postoperative UTIs.

### Comparison with Existing Literature

To the best of our knowledge, this is the first systematic review and meta-analysis on this topic. Our findings for SSIs are consistent with current gynecologic guidelines,^9^ as well as recommendations by the Society for Gynecologic Surgeons Systematic Review Group for vaginal irrigation before vaginal hysterectomy.^30^ Our findings differ, however, from those of a recent meta-analysis in an obstetric population that observed a higher risk of postoperative wound infections and fever in patients who received CHG than PI antisepsis.^31^ This difference could be attributed to several factors. First, the level of contamination and the nature of the surgical procedure itself could impact the antiseptic effect on SSI risk.^32^ Additionally, the vaginal microbiota of pregnant/laboring patients undergoing these procedures may be different.^30,33^

In contrast to our null findings for SSIs, we found a lower pooled incidence of UTIs with use of PI than CHG as a vaginal preparation. The underlying explanation for this finding is unclear but could be related to the greater bactericidal efficacy of PI than CHG against anaerobic organisms and gram-negative bacteria, the leading causes of UTI.^34–37^ Although CHG has been shown to lower total bacterial counts,^38^ studies suggest that it is less bactericidal against anaerobic organisms and gram negative bacteria than against other bacteria. For instance, in a small experimental study, women who performed daily vaginal douching with 0.5% CHG for 1 week had an increased prevalence of gram-negative rods in their flora after 7 days of CHG use than before use.^39^ In addition to differences in bactericidal activity, further possible explanations for our observed lower risk of UTIs with PI than CHG include variation in the formulation concentration, irrigation time, and preparation procedure for CHG across studies,^10^ as no guidance exists regarding the optimal concentration and penetration time of CHG in the vagina (4% vs 2% vs diluted by surgeon).

Lastly, this study did not find statistically significant differences in vaginal irritation between CHG and PI antisepsis. Previous studies have found similar results in patients undergoing cesarean delivery and minor gynecologic surgery.^21,40^ In our systematic review, individual study findings for vaginal irritation were highly heterogeneous, potentially because of the subjective nature of this patient-reported outcome, which might make it more difficult to quantify and report. The high heterogeneity could also be a reflection of the wide range of outcome definitions and scales used across studies. Therefore, future studies should use well-established and consistent scales to assess vaginal irritation.

### Strengths and Limitations

This study has some limitations. 1) Inclusion of retrospective cohort and before and after studies may have introduced confounding by important clinical factors and complications, such as vaginal desquamation by CHG. In addition, the before and after studies implemented additional quality improvement measures (e.g., educational lectures on SSI prevention) at the same time as CHG implementation. This could have led to a change in overall behavior and increased adherence to SSI prevention protocols for patients who received CHG antisepsis. We attempted to mitigate this concern by performing subgroup analyses with only RCTs. 2) Our results were heavily dominated by one retrospective cohort study.^22^ However, that study performed a propensity score matched analysis and had similar baseline characteristics in both vaginal antisepsis groups following matching. 3) Some outcomes were self-reported by patients (vaginal irritation) or may have been missed in participants lost to follow-up, potentially contributing to non-differential outcome misclassification and a bias towards the null. Variability in the vaginal preparation protocols used across studies may have also contributed to our null findings.^20^ Future well-designed studies with larger sample sizes, more comprehensive data collection, and designs or analyses addressing potential confounding factors would be beneficial and may further refine the point estimates and/or direction of our findings. In the meantime, we believe these findings can help guide surgeons in their choice of vaginal antisepsis.

Future research could expand on our findings by comparing the efficacy of different concentrations of CHG and use in specific surgeries (hysterectomy only vs urogynecology procedures, etc.). In addition, assessing the impact of CHG on patient outcomes would provide important information on the overall benefits, risks, and limitations of this antiseptic preparation. Future studies would also benefit from the use of established scales to assess outcomes such as vaginal irritation. This would allow for a more standardized and reliable evaluation of the effects of both interventions on patient outcomes.

## Conclusions and Implications

Vaginal antiseptic solutions reduce infectious morbidity after major gynecologic surgeries. The results of our systematic review suggest that either CHG or PI vaginal preparations can be used before gynecological procedures to prevent SSI, but future randomized studies are warranted to explore the potential for PI to lower postoperative UTI risk.

## Data Availability

All data produced in the present work are contained in the manuscript

## ABBREVIATIONS

ACOG: American College of Obstetricians and Gynecology
CHG: Chlorhexidine Gluconate
CI: Confidence interval
CDC: Centers for Disease Control and Prevention
FIGO: International Federation of Gynecology and Obstetrics
PI: Povidone Iodine
PSM: Propensity score matching
PRISMA: Preferred Reporting Items for Systematic Reviews and Meta-analysis
PROSPERO: International Prospective Register of Systematic Reviews
RR: Relative Risk
ROBINS-I: Risk of bias in non-randomized studies of interventions
RoB 2: Risk of bias in randomized trials
RCT: Randomized Controlled Trial
SSI: Surgical Site Infection
UTI: Urinary Tract Infection

## Condensation page

**Tweetable statement:** Comparing vaginal preparation for urogynecological surgeries using Povidone Iodine versus Chlorhexidine Gluconate: A Systematic Review and Meta-Analysis

**Short Title:** Povidone Iodine vs Chlorhexidine vaginal antisepsi**s**

**AJOG at a Glance:** Although vaginal antisepsis is recommended before gynecological surgery, there is no consensus over the preferred vaginal preparation solution. We aimed to perform a meta-analysis of post-surgical outcomes comparing Povidone Iodine (PI) versus Chlorhexidine Gluconate (CHG) for vaginal antisepsis of urogynecological procedures to assess for incremental benefits. The results of our study shows that vaginal antisepsis with PI is associated with a lower incidence of UTIs post urogynecological operations and the use of PI or CHG most likely have no clinical significance for SSIs. Our findings support current guidelines that either form of vaginal antisepsis can be used.

## Appendix 1

### Full Search Strategies

#### Embase

Date Searched: 12/15/2023

Applied Database Supplied Limits: none

Number of Results: 353

##### Full Search Strategy

(‘gynecologic surgery’/exp OR ‘vagina surgery’/exp OR ‘hysterectomy’/exp OR ‘vaginal hysterectomy’/exp OR ‘total hysterectomy’/exp OR ‘urogynecologic surgery’/exp OR ((gynecologic OR gynaecologic OR gynaecological OR urogynecologic OR vagina OR vaginal) NEAR/3 (surger* OR surgical OR operation* OR operative)):ti,ab,kw OR (hysterectomy OR ‘supervaginal amputation’ OR ‘uterus amputation’ OR ‘uterus extirpation’ OR Hysterectomy OR panhysterectomy):ti,ab,kw) AND (‘chlorhexidine’/exp OR ‘chlorhexidine gluconate’/exp OR (alcloxidine OR bactoscrub OR ‘bactosept concentrate’ OR bidex OR bioscrub OR bucoglobin OR chlorhex OR chlorhexamed OR chlorhexidin OR Chlorhexidine OR chlorhexidine OR chlorhexidinium OR chlorohex OR chlorohexydine OR ‘cida stat’ OR cidastat OR ‘cida-stat’ OR cleardent OR clohexidine OR clorhexidine OR corsodyl OR dosiseptine OR ‘dyna-hex’ OR ‘e z scrub’ OR exidine OR exitane OR ‘e-z scrub’ OR fectin OR hexadent OR hibiclens OR hibident OR hibidil OR hibigel OR hibiguard oR hibiprep OR hibiscrub OR hibisol OR hibistat OR hibitan OR hibitane OR hidine OR ‘improved phisohex’ OR klorheksidos OR klorhexidin OR klorhexol OR ‘lemocin cx’ OR lisium OR nibitane OR nolvasan OR nolvascin OR peridex OR ‘perio chip’ OR periochip OR periodentix OR periogard OR perioxidin OR perisol OR plakout OR plurexid OR ‘prevacare r’ OR ‘readyprep chg’ OR rotersept OR savacol OR septalone OR septol OR sterilon OR ‘steri-stat’ OR tubilicid OR tubulicid OR umbipro):ti,ab,kw OR ‘povidone iodine’/exp OR ‘iodine’/exp OR (‘beta isodona’ OR betadine OR betaisodona OR braunoderm OR braunol OR braunovidon OR bridina OR bridine OR destrobac OR iodine OR iodium OR iodopovidone OR iosal OR isodine OR jodium OR medadine OR ‘neo hydriol fluid’ OR ‘neo hydriol viscous’ OR pharmadine OR povadyne OR ‘povidone-iodine’ OR prepodyne OR proviodine OR traumasept OR videne OR vidine):ti,ab,kw) AND (‘adverse event’/exp OR ‘procedural site reaction’/exp OR ‘cellulitis’/exp OR ‘irritation’/exp OR ‘morbidity’/exp OR ‘outcome assessment’/exp OR ‘hospital readmission’/exp OR ‘postoperative complication’/exp OR ‘postoperative infection’/exp OR ‘reoperation’/exp OR ‘sepsis’/exp OR ‘surgical infection’/exp OR ‘urinary tract infection’/exp OR ‘vaginal irritation’/exp OR ‘vaginitis’/exp OR ‘wound infection’/exp OR (cellulitis OR irritation OR morbidity OR readmission* OR rehospitalization* OR reoperation* OR sepsis OR ‘septic disease’ OR ‘UTI’ OR ‘UTIs’ OR vaginitis OR colpitis OR kolpitis OR ‘vagina inflammation’) OR (adverse NEAR/3 (event* OR reaction* OR effect*)):ti,ab,kw OR (outcome* NEAR/2 (assessment* OR measurement* OR surgical OR surgery)):ti,ab,kw OR ((postoperative OR postoperation OR ‘post-operation’ OR ‘post-operative’ OR ‘after operation’ OR ‘post-surgery’ OR ‘post-surgical’ OR postsurg* OR surger* OR ‘surgery-associated’ OR ‘surgery-derived’ OR ‘surgery-induced’ OR ‘surgery-related’) NEAR/3 (complication* OR infection*)):ti,ab,kw OR ((‘surgical site’ OR wound OR urinary OR ‘urine tract’ OR urologic*) NEAR/3 (infected OR infection* OR contaminated OR contamination OR sepsis)):ti,ab,kw)

#### Ovid Medline

Date Searched: 12/15/2023

Applied Database Supplied Limits: none

Number of Results: 142

##### Full Search Strategy

(exp Gynecologic Surgical Procedures/ OR exp Hysterectomy, Vaginal/ OR exp Hysterectomy/ OR ((gynecologic OR gynaecologic OR gynaecological OR urogynecologic OR vagina OR vaginal) ADJ3 (surger* OR surgical OR operation* OR operative)).ti,ab,kf. OR (hysterectomy OR supervaginal amputation OR uterus amputation OR uterus extirpation OR Hysterectomy OR panhysterectomy).ti,ab,kf.) AND (exp Chlorhexidine/ OR (alcloxidine OR bactoscrub OR bactosept concentrate OR bidex OR bioscrub OR bucoglobin OR chlorhex OR chlorhexamed OR chlorhexidin OR Chlorhexidine OR chlorhexidine OR chlorhexidinium OR chlorohex OR chlorohexydine OR cida stat OR cidastat OR cida-stat OR cleardent OR clohexidine OR clorhexidine OR corsodyl OR dosiseptine OR dyna-hex OR e z scrub OR exidine OR exitane OR e-z scrub OR fectin OR hexadent OR hibiclens OR hibident OR hibidil OR hibigel OR hibiguard oR hibiprep OR hibiscrub OR hibisol OR hibistat OR hibitan OR hibitane OR hidine OR improved phisohex OR klorheksidos OR klorhexidin OR klorhexol OR lemocin cx OR lisium OR nibitane OR nolvasan OR nolvascin OR peridex OR perio chip OR periochip OR periodentix OR periogard OR perioxidin OR perisol OR plakout OR plurexid OR prevacare r OR readyprep chg OR rotersept OR savacol OR septalone OR septol OR sterilon OR steri-stat OR tubilicid OR tubulicid OR umbipro).ti,ab,kf. OR exp Iodine/ OR exp Povidone-Iodine/ OR (beta isodona OR betadine OR betaisodona OR braunoderm OR braunol OR braunovidon OR bridina OR bridine OR destrobac OR iodine OR iodium OR iodopovidone OR iosal OR isodine OR jodium OR medadine OR neo hydriol fluid OR neo hydriol viscous OR pharmadine OR povadyne OR povidone-iodine OR prepodyne OR proviodine OR traumasept OR videne OR vidine).ti,ab,kf.) AND (exp Cellulitis/ OR exp Morbidity/ OR exp Outcome Assessment, Health Care/ OR exp Patient Readmission/ OR exp Postoperative Complications/ OR exp Surgical Wound Infection/ OR exp Postoperative Complications/ OR exp Reoperation/ OR exp Sepsis/ OR exp Urinary Tract Infections/ OR exp Vaginitis/ OR exp Wound Infection/ OR (cellulitis OR irritation OR morbidity OR readmission* OR rehospitalization* OR reoperation* OR sepsis OR septic disease OR UTI OR UTIs OR vaginitis OR colpitis OR kolpitis OR vagina inflammation) OR (adverse ADJ3 (event* OR reaction* OR effect*)).ti,ab,kf. OR (outcome* ADJ2 (assessment* OR measurement* OR surgical OR surgery)).ti,ab,kf. OR ((postoperative OR postoperation OR post-operation OR post-operative OR after operation OR post-surgery OR post-surgical OR postsurg* OR surger* OR surgery-associated OR surgery-derived OR surgery-induced OR surgery-related) ADJ3 (complication* OR infection*)).ti,ab,kf. OR ((surgical site OR wound OR urinary OR urine tract OR urologic*) ADJ3 (infected OR infection* OR contaminated OR contamination OR sepsis)).ti,ab,kf.)

#### Scopus

Date Searched: 12/15/2023

Applied Database Supplied Limits: none

Number of Results: 291

##### Full Search Strategy

((TITLE-ABS-KEY((gynecologic OR gynaecologic OR gynaecological OR urogynecologic OR vagina OR vaginal) W/3 (surger* OR surgical OR operation* OR operative))) OR (TITLE-ABS-KEY(hysterectomy OR “supervaginal amputation” OR “uterus amputation” OR “uterus extirpation” OR Hysterectomy OR panhysterectomy))) AND ((TITLE-ABS-KEY(alcloxidine OR bactoscrub OR “bactosept concentrate” OR bidex OR bioscrub OR bucoglobin OR chlorhex OR chlorhexamed OR chlorhexidin OR Chlorhexidine OR chlorhexidine OR chlorhexidinium OR chlorohex OR chlorohexydine OR “cida stat” OR cidastat OR “cida-stat” OR cleardent OR clohexidine OR clorhexidine OR corsodyl OR dosiseptine OR “dyna-hex” OR “e z scrub” OR exidine OR exitane OR “e-z scrub” OR fectin OR hexadent OR hibiclens OR hibident OR hibidil OR hibigel OR hibiguard oR hibiprep OR hibiscrub OR hibisol OR hibistat OR hibitan OR hibitane OR hidine OR “improved phisohex” OR klorheksidos OR klorhexidin OR klorhexol OR “lemocin cx” OR lisium OR nibitane OR nolvasan OR nolvascin OR peridex OR “perio chip” OR periochip OR periodentix OR periogard OR perioxidin OR perisol OR plakout OR plurexid OR “prevacare r” OR “readyprep chg” OR rotersept OR savacol OR septalone OR septol OR sterilon OR “steri-stat” OR tubilicid OR tubulicid OR umbipro)) OR (TITLE-ABS-KEY(“beta isodona” OR betadine OR betaisodona OR braunoderm OR braunol OR braunovidon OR bridina OR bridine OR destrobac OR iodine OR iodium OR iodopovidone OR iosal OR isodine OR jodium OR medadine OR “neo hydriol fluid” OR “neo hydriol viscous” OR pharmadine OR povadyne OR “povidone-iodine” OR prepodyne OR proviodine OR traumasept OR videne OR vidine))) AND ((TITLE-ABS-KEY(cellulitis OR irritation OR morbidity OR readmission* OR rehospitalization* OR reoperation* OR sepsis OR “septic disease” OR “UTI” OR “UTIs” OR vaginitis OR colpitis OR kolpitis OR “vagina inflammation”)) OR (TITLE-ABS-KEY(adverse W/3 (event* OR reaction* OR effect*))) OR (TITLE-ABS-KEY(outcome* W/2 (assessment* OR measurement* OR surgical OR surgery))) OR (TITLE-ABS-KEY((postoperative OR postoperation OR “post-operation” OR “post-operative” OR “after operation” OR “post-surgery” OR “post-surgical” OR postsurg* OR surger* OR “surgery-associated” OR “surgery-derived” OR “surgery-induced” OR “surgery-related”) W/3 (complication* OR infection*))) OR (TITLE-ABS-KEY((“surgical site” OR wound OR urinary OR “urine tract” OR urologic*) W/3 (infected OR infection* OR contaminated OR contamination OR sepsis))))

#### The Cochrane Library

Date Searched: 12/15/2023

Applied Database Supplied Limits: none

Number of Results

CENTRAL: 103

CDSR: 1

##### Full Search Strategy

([mh “Gynecologic Surgical Procedures”] OR [mh “Hysterectomy, Vaginal”] OR [mh “Hysterectomy”] OR ((gynecologic OR gynaecologic OR gynaecological OR urogynecologic OR vagina OR vaginal) NEAR/3 (surger* OR surgical OR operation* OR operative)):ti,ab,kw OR (hysterectomy OR “supervaginal amputation” OR “uterus amputation” OR “uterus extirpation” OR Hysterectomy OR panhysterectomy):ti,ab,kw) AND ([mh “Chlorhexidine”] OR (alcloxidine OR bactoscrub OR bactosept concentrate OR bidex OR bioscrub OR bucoglobin OR chlorhex OR chlorhexamed OR chlorhexidin OR Chlorhexidine OR chlorhexidine OR chlorhexidinium OR chlorohex OR chlorohexydine OR “cida stat” OR cidastat OR “cida stat” OR cleardent OR clohexidine OR clorhexidine OR corsodyl OR dosiseptine OR “dyna hex” OR “e z scrub” OR exidine OR exitane OR “e z scrub” OR fectin OR hexadent OR hibiclens OR hibident OR hibidil OR hibigel OR hibiguard oR hibiprep OR hibiscrub OR hibisol OR hibistat OR hibitan OR hibitane OR hidine OR “improved phisohex” OR klorheksidos OR klorhexidin OR klorhexol OR “lemocin cx” OR lisium OR nibitane OR nolvasan OR nolvascin OR peridex OR “perio chip” OR periochip OR periodentix OR periogard OR perioxidin OR perisol OR plakout OR plurexid OR “prevacare r” OR “readyprep chg” OR rotersept OR savacol OR septalone OR septol OR sterilon OR “steri stat” OR tubilicid OR tubulicid OR umbipro):ti,ab,kw OR [mh “Iodine”] OR [mh “Povidone-Iodine”] OR (“beta isodona” OR betadine OR betaisodona OR braunoderm OR braunol OR braunovidon OR bridina OR bridine OR destrobac OR iodine OR iodium OR iodopovidone OR iosal OR isodine OR jodium OR medadine OR “neo hydriol fluid” OR “neo hydriol viscous” OR pharmadine OR povadyne OR “povidone iodine” OR prepodyne OR proviodine OR traumasept OR videne OR vidine):ti,ab,kw) AND ([mh “Cellulitis”] OR [mh “Morbidity”] OR [mh “Outcome Assessment, Health Care”] OR [mh “Patient Readmission”] OR [mh “Postoperative Complications”] OR [mh “Surgical Wound Infection”] OR [mh “Postoperative Complications”] OR [mh “Reoperation”] OR [mh “Sepsis”] OR [mh “Urinary Tract Infections”] OR [mh “Vaginitis”] OR [mh “Wound Infection”] OR (cellulitis OR irritation OR morbidity OR readmission* OR rehospitalization* OR reoperation* OR sepsis OR “septic disease” OR “UTI” OR “UTIs” OR vaginitis OR colpitis OR kolpitis OR “vagina inflammation”) OR (adverse NEAR/3 (event* OR reaction* OR effect*)):ti,ab,kw OR (outcome* NEAR/2 (assessment* OR measurement* OR surgical OR surgery)):ti,ab,kw OR ((postoperative OR postoperation OR “post operation” OR “post operative” OR “after operation” OR “post surgery” OR “post surgical” OR postsurg* OR surger* OR “surgery associated” OR “surgery derived” OR “surgery induced” OR “surgery related”) NEAR/3 (complication* OR infection*)):ti,ab,kw OR ((“surgical site” OR wound OR urinary OR “urine tract” OR urologic*) NEAR/3 (infected OR infection* OR contaminated OR contamination OR sepsis)):ti,ab,kw)

#### ClinicalTrials.gov

Date Searched: 12/15/2023

Number of Results: 2

##### Full Search Strategy

*“gynecologic surgeries” -other terms*

AND

(Chlorhexidine OR Iodine) -*intervention/treatment*

AND

(“Surgical Site Infection” OR “urinary tract infections” OR “Vaginal Irritation” OR sepsis OR morbidity) - *condition/disease*

## Appendix 2: Table of eligibility criteria

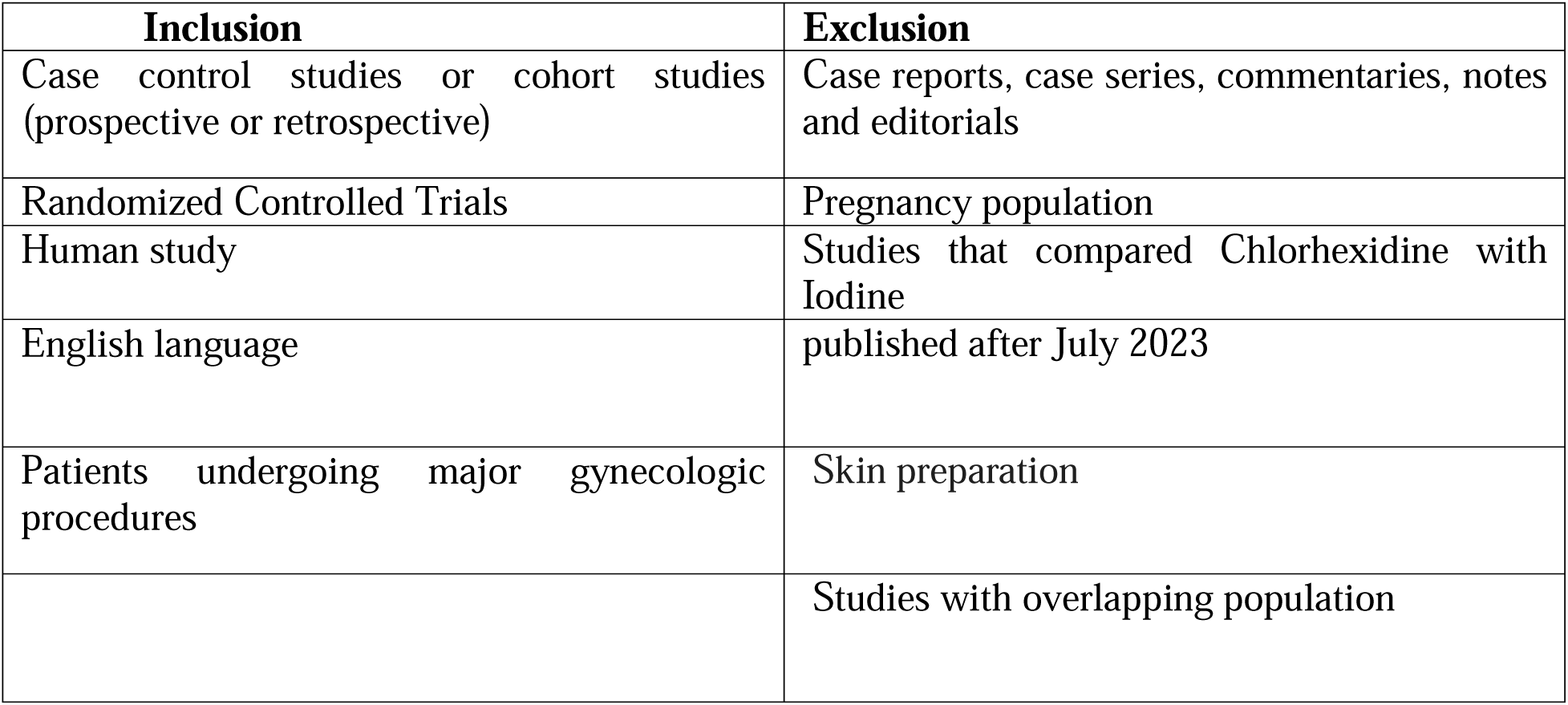

| Inclusion | Exclusion |
| --- | --- |
| Case control studies or cohort studies (prospective or retrospective) | Case reports, case series, commentaries, notes and editorials |
| Randomized Controlled Trials | Pregnancy population |
| Human study | Studies that compared Chlorhexidine with Iodine |
| English language | published after July 2023 |
| Patients undergoing major gynecologic procedures | Skin preparation |
|  | Studies with overlapping population |

## SUPPLEMENTARY APPENDICES AND TABLE

**Table S1:** Risk of bias appraisal of non-randomized studies using ROBIN-I tool.

|  |  | Risk of bias domains |  |  |  |  |  |  |  |
| --- | --- | --- | --- | --- | --- | --- | --- | --- | --- |
|  |  | D1 | D2 | D3 | D4 | D5 | D6 | D7 | Overall |
| Study | Al-Niami 2016 | ! | + | + | + | - | - | X | ! |
|  | Schwartz 2019 | X | - | X | ? | ? | X | X | X |
|  | Skeith 2021 | - | - | - | + | - | - | - | - |
|  | Wong 2021 | X | - | - | ? | ? | - | X | X |

Domains:
D1: Bias due to confounding. D2: Bias due to selection of participants. D3: Bias in classification of interventions. D4: Bias due to deviations from intended interventions. D5: Bias due to missing data. D6: Bias in measurement of outcomes. D7: Bias in selection of the reported result.
Judgement
Critical
Serious
Moderate
Low
No information

**Figure S1:**
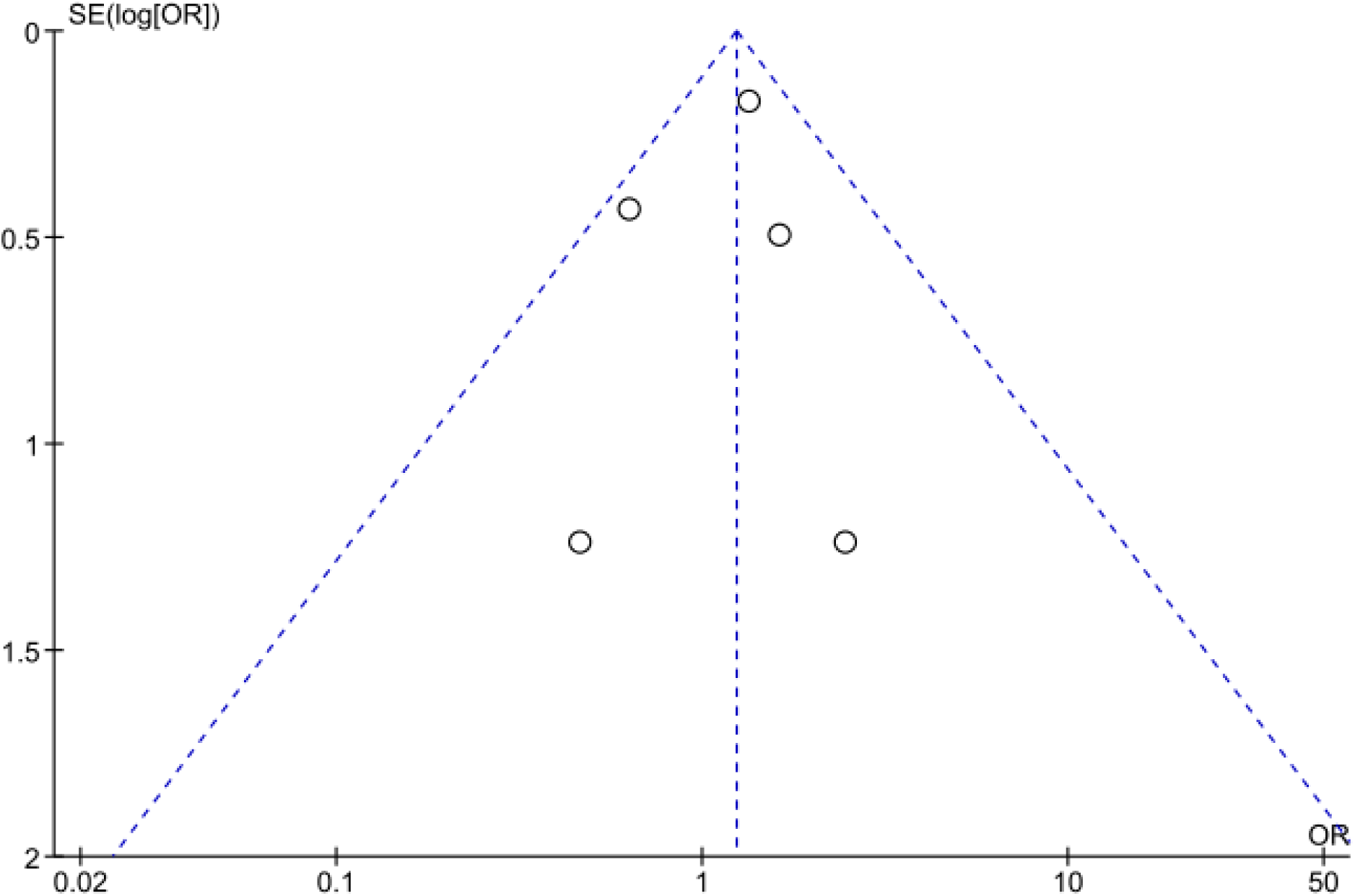
Funnel plot for surgical site infections (SSIs).

